# Identifying main and interaction effects of risk factors to predict intensive care admission in patients hospitalized with COVID-19: a retrospective cohort study in Hong Kong

**DOI:** 10.1101/2020.06.30.20143651

**Authors:** Jiandong Zhou, Gary Tse, Sharen Lee, Tong Liu, William KK Wu, Zhidong Cao, Daniel Dajun Zeng, Ian Chi Kei Wong, Qingpeng Zhang, Bernard Man Yung Cheung

## Abstract

**Background:** The coronavirus disease 2019 (COVID-19) has become a pandemic, placing significant burdens on the healthcare systems. In this study, we tested the hypothesis that a machine learning approach incorporating hidden nonlinear interactions can improve prediction for Intensive care unit (ICU) admission.

**Methods:** Consecutive patients admitted to public hospitals between 1^st^ January and 24^th^ May 2020 in Hong Kong with COVID-19 diagnosed by RT-PCR were included. The primary endpoint was ICU admission.

**Results:** This study included 1043 patients (median age 35 (IQR: 32-37; 54% male). Nineteen patients were admitted to ICU (median hospital length of stay (LOS): 30 days, median ICU LOS: 16 days). ICU patients were more likely to be prescribed angiotensin converting enzyme inhibitors/angiotensin receptor blockers, anti-retroviral drugs lopinavir/ritonavir and remdesivir, ribavirin, steroids, interferon-beta and hydroxychloroquine. Significant predictors of ICU admission were older age, male sex, prior coronary artery disease, respiratory diseases, diabetes, hypertension and chronic kidney disease, and activated partial thromboplastin time, red cell count, white cell count, albumin and serum sodium. A tree-based machine learning model identified most informative characteristics and hidden interactions that can predict ICU admission. These were: low red cells with 1) male, 2) older age, 3) low albumin, 4) low sodium or 5) prolonged APTT. A five-fold cross validation confirms superior performance of this model over baseline models including XGBoost, LightGBM, random forests, and multivariate logistic regression.

**Conclusions:** A machine learning model including baseline risk factors and their hidden interactions can accurately predict ICU admission in COVID-19.

## Introduction

Coronavirus disease 2019 (COVID-19), the third coronavirus epidemic in the recent two decades after severe acute respiratory syndrome (SARS) and Middle East respiratory syndrome (MERS), has become a pandemic, placing significant burdens on healthcare systems worldwide ^1^. The number of people confirmed with COVID-19 worldwide exceeded 7.4 million on June 11, 2020, including at least 416,000 deaths across 188 countries and territories ^2^. The coronavirus pandemic remains unresolved, even though countries around the world have moved to lift quarantines, stay-at-home orders and other social restrictions. A particular challenge countries face in the COVID-19 pandemic is the surge in demand for intensive care unit (ICU) care ^3, 4^. Recent studies have exposed an astonishing case fatality rate of 61.5% for critical cases, increasing sharply with older age and for patients with underlying comorbidities ^5^. The unfulfilled ICU demand would immediately lead to elevated fatality rate. The critical question on the clinical characteristics and relevant biomarkers for efficient ICU management of COVID-19 patients remains unanswered ^6^. Identification of prognostic biomarkers to distinguish patients that require immediate medical attention has become an urgent yet challenging necessity. Therefore, the aim of this study is to identify significant risk factors or characteristics as well as hidden interaction effects associated with ICU admission by using an interpretable machine learning approach.

## Methods

### Study design and population

This study was approved by the Institutional Review Board of the University of Hong Kong/Hospital Authority Hong Kong West Cluster. This was a retrospective, territory-wide cohort study of patients infected with COVID-19, as confirmed by RT-PCR, between 1^st^ January and 24^th^ May 2020. The patients were identified from the Clinical Data Analysis and Reporting System (CDARS), a territory-wide database that centralizes patient information from individual local hospitals to establish comprehensive medical data, including clinical characteristics, disease diagnosis, laboratory results, and drug treatment details. The system has been previously used by both our team and other teams in Hong Kong ^7-10^. Clinical data include primary diagnoses after admission (1^st^ January 2020 to 24^th^ May 2020) and comorbidities (1^st^ January 1999 to 31^st^ December 2019) in the past decade. The list of conditions identified is detailed in the **Supplementary Appendix**. Diagnosis of COVID-19 was made by RT-PCR. Other respiratory viruses, including influenza A virus (H1N1, H3N2, H7N9), influenza B virus, respiratory syncytial virus, parainfluenza virus, adenovirus, SARS coronavirus (SARS-CoV), and MERS coronavirus (MERS-CoV) were also examined with RT-PCR.

### Outcomes and statistical analysis

The primary outcome was ICU admission. Continuous variables were presented as median (95% confidence interval [CI] or interquartile range [IQR]) and categorical variables were presented count (%). The Mann-Whitney U test was used to compare continuous variables. The χ^2^ test with Yates’ correction was used for 2×2 contingency data, and Pearson’s χ^2^ test was used for contingency data for variables with more than two categories. To identify the significant risk factors associated with ICU admission of COVID-19 patients, univariate logistic regression was used to estimate odds ratios (ORs) and 95% CIs, adjusting for age, sex, comorbidities. A two-sided α of less than 0.05 was considered statistically significant. Statistical analyses (including univariate logistic regression) were performed using RStudio software (Version: 1.1.456) and Python (Version: 3.6).

### Development of a tree-based interpretable machine learning model

After the identification of significant predictors for ICU admission, we aim to further construct a practically useful ICU use decision-making model by considering both main and interaction effects among those important univariable variables. Here the interaction effects, mainly pairwise interactions, capture the hidden nonlinear dependence between risk characteristics and can provide additional information for ICU outcome identification, besides individual predictors. Significant predictors identified on univariate logistic regression were enter into a state-of-the-art interpretable boosting machine model: Explainable Boosting Machine (EBM) ^11^.

The EBM model is an explainable supervised predictor developed by using modern machine learning techniques like bagging, gradient boosting, and automatic main and interaction effects detection with high accuracy of state-of-the-art learning models (e.g., random forests ^12^ and XGBoost ^13^) with its light memory usage and fast prediction time. EBM is constructed with multiple hierarchically organized simple classifiers consisting of sequences of binary decisions. Unlike these black-box models, EBM produce lossless explanations for outcome predictions due to its great interpretability potential of tree-based decision system, which is desired for clinically operable decision-making. In contrast, internally black-box-like learning models are typically difficult to interpret. Intrinsic interpretability as equipped in EBM aims to intrinsically interpret the model predictions. The contribution of main and interaction effects to identify ICU use can be determined by their accumulated use in each decision tree splitting process, which can be easily sorted and visualized in descending order to identify the more important variables.

## Results

### Baseline characteristics

The flowchart of patient enrolment in this study is provided in **Figure 1**. A total of 1043 patients admitted to the hospital between 1^st^ January 2020 and 24^th^ May 2020 were included in this study. The case distributions with respect to the different districts of Hong Kong are shown in **Figure 2**. There are 373 cases from Hong Kong Island District (36%), 212 cases from Kowloon District (20%), 398 cases from New Territories District (38%), and 60 cases without district indicators (6%). Chinese is the most common nationality (914, 87.6%), followed by Filipino (38, 3.6%), Pakistani (22, 2.1%), British (14, 1.3%), French (9, 0.9%), Nepalese (9, 0.9%), American (6, 0.58%), Indian (4, %), Canadian (2, 0.2%), Australian (2, 0.2%) and Korean, German, New Zealander, Greek, Thai, Indonesian, Japanese, and Netherlander (1 each, 0.1%).

**Figure 1.**
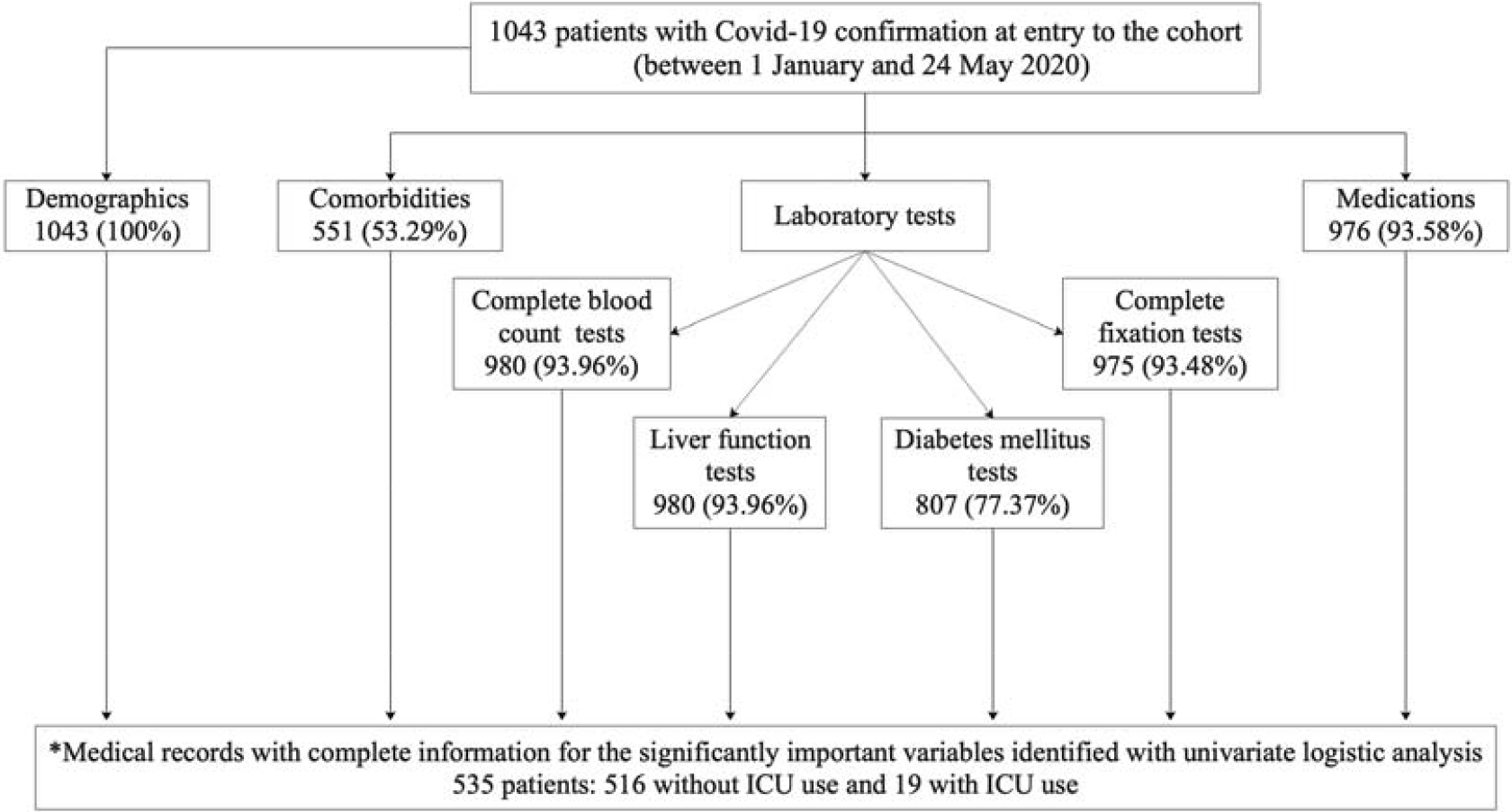
Flowchart of patient enrolment. * denotes the imposition of multiple criteria; Study baseline was defined as 24 hours after arrival at hospital; COVID-19 = coronavirus disease 2019; ICU=intensive care unit.

**Figure 2.**
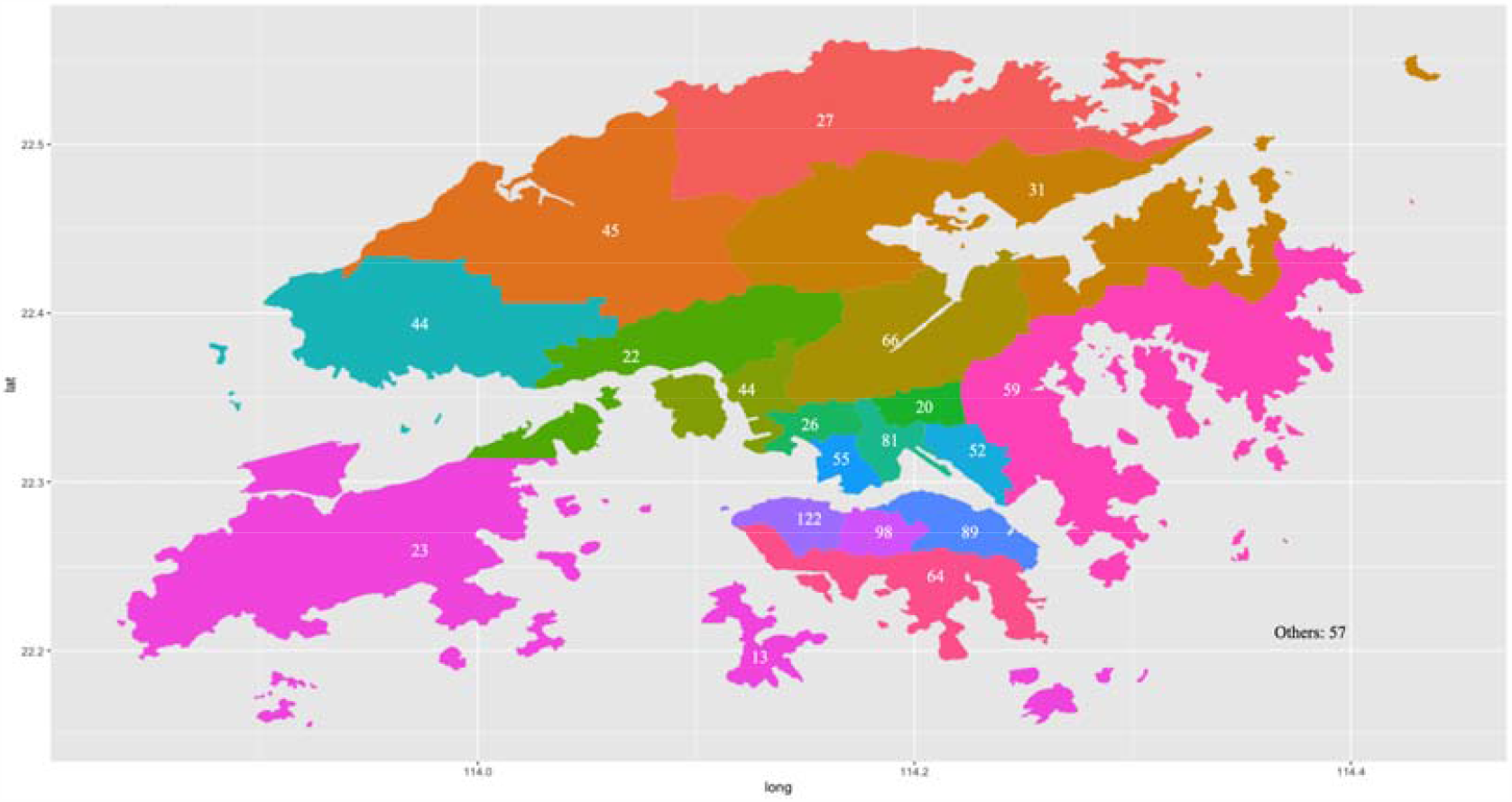
Distribution of COVID-19 patients in Hong Kong districts of residence

The baseline demographics, comorbidities, medications, and laboratory test findings are shown in **Table 1**. Of the included patients, 563 were males (54%, median age: 35 [IQR: 32-37], range: 0-93 years old) and 480 were females (46%, median age: 35 [IQR: 32-37], range 1-96 years old) (**Figure 3**). Most patients (n=776, 74%) were between 18 and 60 years of age. In total, 19 patients (14 males, 73.68%, median ICU length of stay [LOS]: 16 days) were admitted to the ICU and the numbers within age intervals are also shown in **Figure 3**. Patients admitted to the ICU has median inpatient length of stay (LOS) of 30 days, in comparison to a median of 20 days for patients without ICU admission. The timeline of COVID-19 cases after hospitalization is shown in **Figure 4**. Amongst the ICU patients, there are 12 Chinese, 1 Filipino, and 6 of unknown race. The 19 ICU patients has median urine output of 1610 ml/24 hours (IQR: 1255-2000, max: 3310). The distributions of other physiological parameters are shown in **Table 1**. In addition, 9 ICU patients (47.37%) were on ventilators for respiratory support, and three (15.79%) received renal replacement therapy. Four patients died. Two deaths occurred during inpatient hospitalization with single admission, one during ICU hospitalization, and one upon admission.

**Table 1.** Physiological parameter distributions of patients with ICU admission

| Parameter | High: Median (IQR, max) | Low: Median (IQR, max) |
| --- | --- | --- |
| White cell count, x 10<sup>9</sup>/L | 8.11 (5.65-13.85, 24.25) | 6 (4.525-9.17, 16.61) |
| Urea, mmol/L | 5.9 (4.5-8.8, 13.8) | - |
| Platelet count, x 10<sup>9</sup>/L | 240 (220.5-318.5, 472) | 235 (202-282.5, 461) |
| Na<sup>+</sup>, mmol/L | 137.3 (135.35-140.45, 147) | 135.6 (133.6-139.95, 145) |
| Mean BP, mmHg | 110 (100-123.5, 183) | 67 (61-74, 93) |
| K<sup>+</sup>, mmol/L | 4.17 (3.675-4.45, 5.3) | 3.66 (3.15-3.95, 4.39) |
| Highest Respiratory Rate, /min | 29 (24.5-32, 44) | 19 (14-20.5, 30) |
| Heart rate, /min | 92 (86-110, 154) | 60 (52.5-70.5, 84) |
| Hematocrit | 0.378 (0.355-0.413, 0.5) | 0.363 (0.308-0.4095, 0.5) |
| Hemoglobin, g/dL | 13 (12.35-14, 16.5) | 12.6 (10.7-13.65, 16.5) |
| Blood glucose, mmol/L | 8.6 (6.85-12.35, 25.8) | 5.9 (4.9-6.85, 10.5) |
| Creat, mmol/L | 73 (57-102.45, 266) | 69 (54.5-86.5, 173.3) |
| Core temperature, °c | 38.3 (37.4-39, 40.8) | 36.6 (36.15-37.05, 38.4) |
| Bilirubin, umol/L | 14 (9-22.1, 79) | - |
| Albumin, g/L | 27.6 (23.5-34.15, 48.4) | 25 (21-30.6, 48.4) |

**Figure 3.**
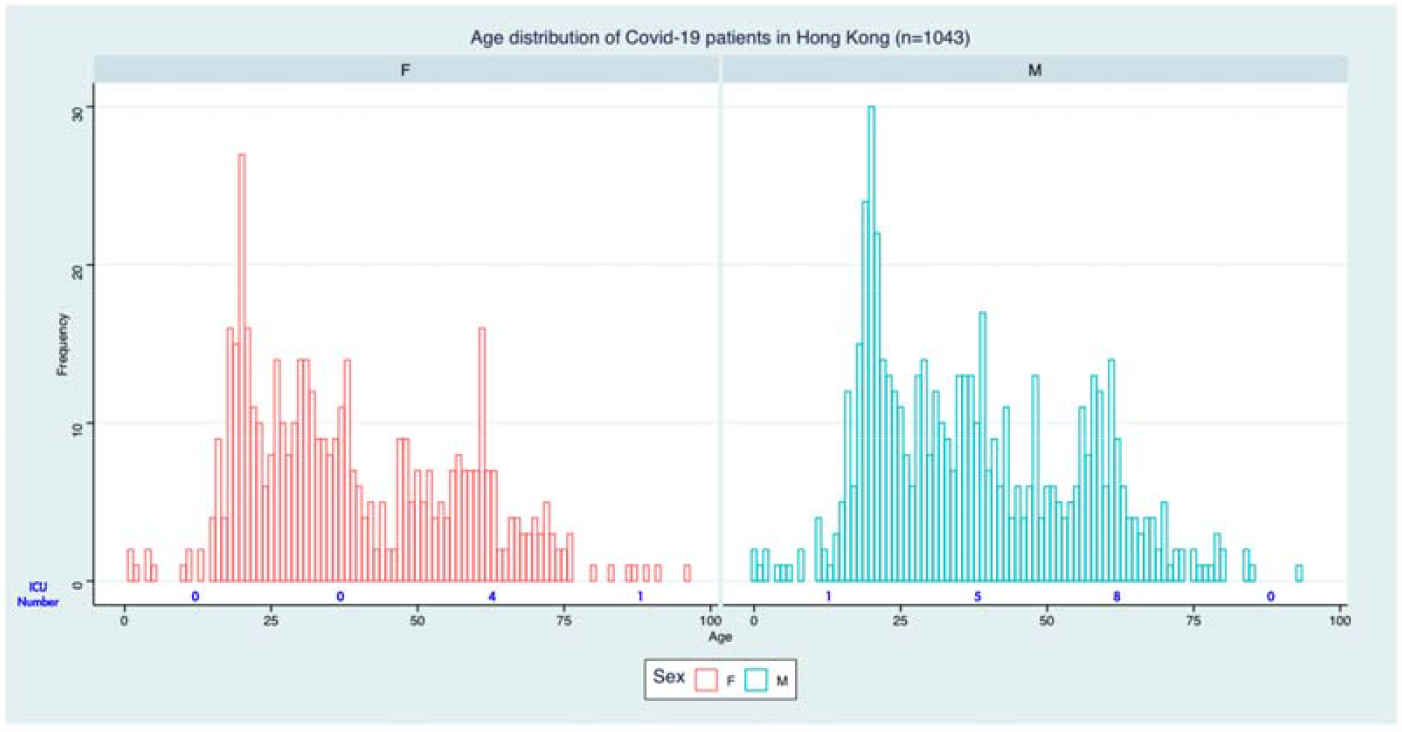
Age distribution of COVID-19 inpatients in Hong Kong.

**Figure 4.**
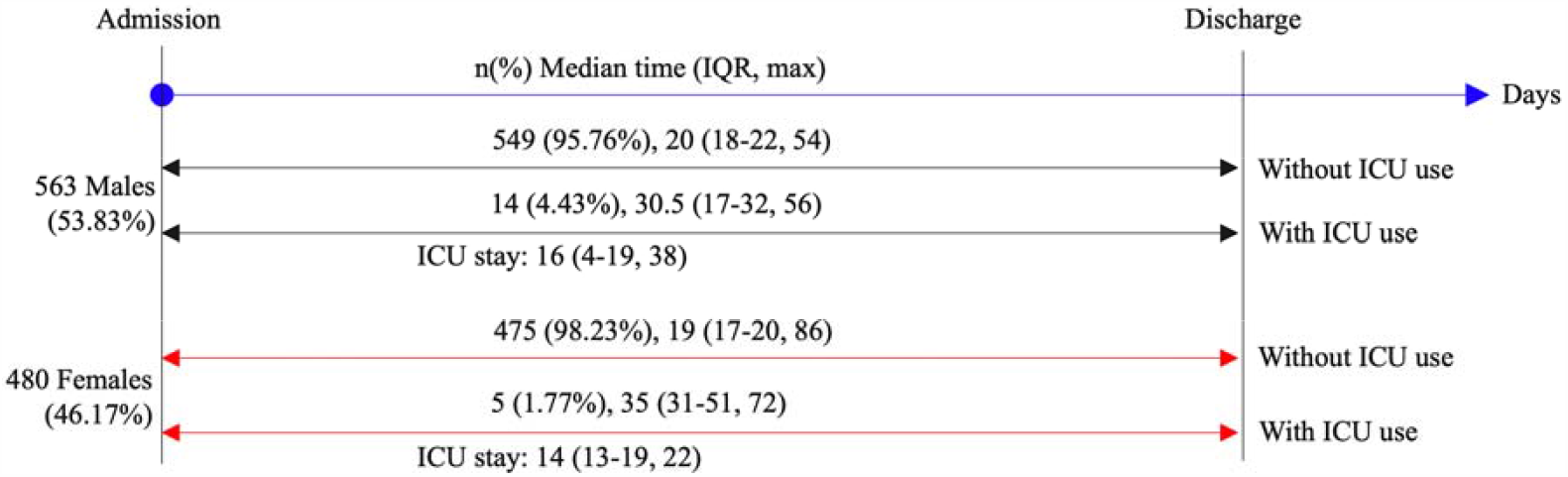
Timeline of COVID-19 cases after hospitalization.

A total of 535 COVID-19 patients (87.3%) had records of preexisting comorbidities (n=1237) between January 1^st^, 1999 to December 31^st^, 2019. Of these, 230 patients (42.99%) had respiratory diseases, 174 patients (32.52%) had gastrointestinal diseases, 108 patients (20.19%) had hypertension, 54 patients (10.09%) had diabetes, 21 patients (3.93%) had chronic kidney diseases, and 10 patients (1.87%) had cardiovascular diseases.

In terms of medications prescribed during the inpatient stay for non-ICU patients, lopinavir/ritonavir (Kaletra) is the most commonly used drug (60.8%), followed by ribavirin (53.2%), interferon-beta (32.5%), angiotensin converting enzyme inhibitors (ACEI) or angiotensin receptor blockers (ARBs) (18.9%), steroids (14.6%), hydroxychloroquine (13.2%), and remdesivir (2.5%). Among the ICU patients, lopinavir/ritonavir was the most frequently prescribed drug (88.9%), followed by ribavirin (77.8%), ACEI/ARB (77.8%), hydroxychloroquine (38.9%), interferon-beta (38.9%), steroids (27.8%) and remdesivir (5.6%). We find that patients admitted to ICU are more likely to be given Kaletra and ribavirin, which may reflect more aggressive treatment towards critically ill patients.

### Predictors of ICU admission

Univariate logistic regression was conducted to identify significant predictors of ICU admission (**Table 3**). The following are significant predictors for ICU admission of COVID-19 patients:

**Table 2.**
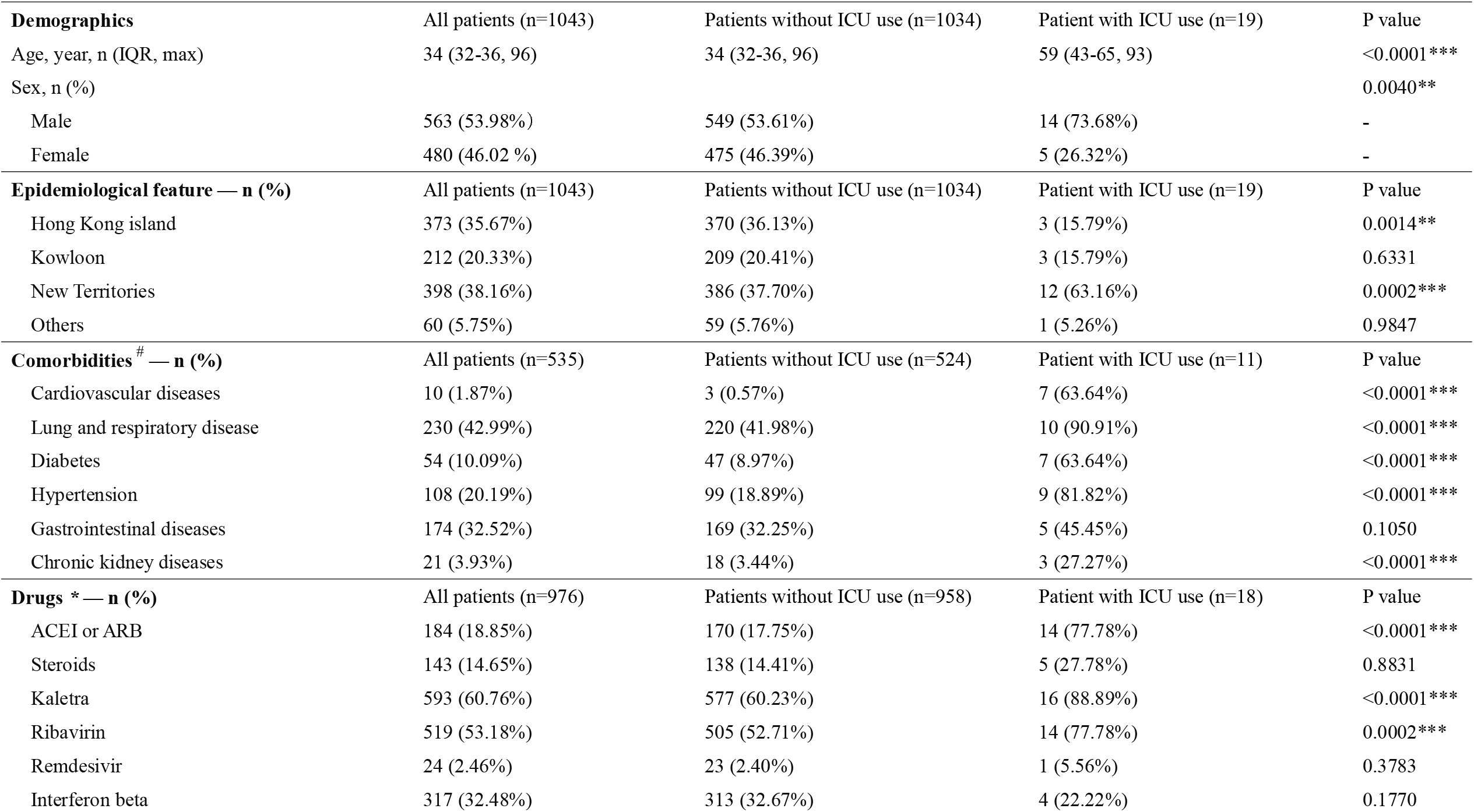
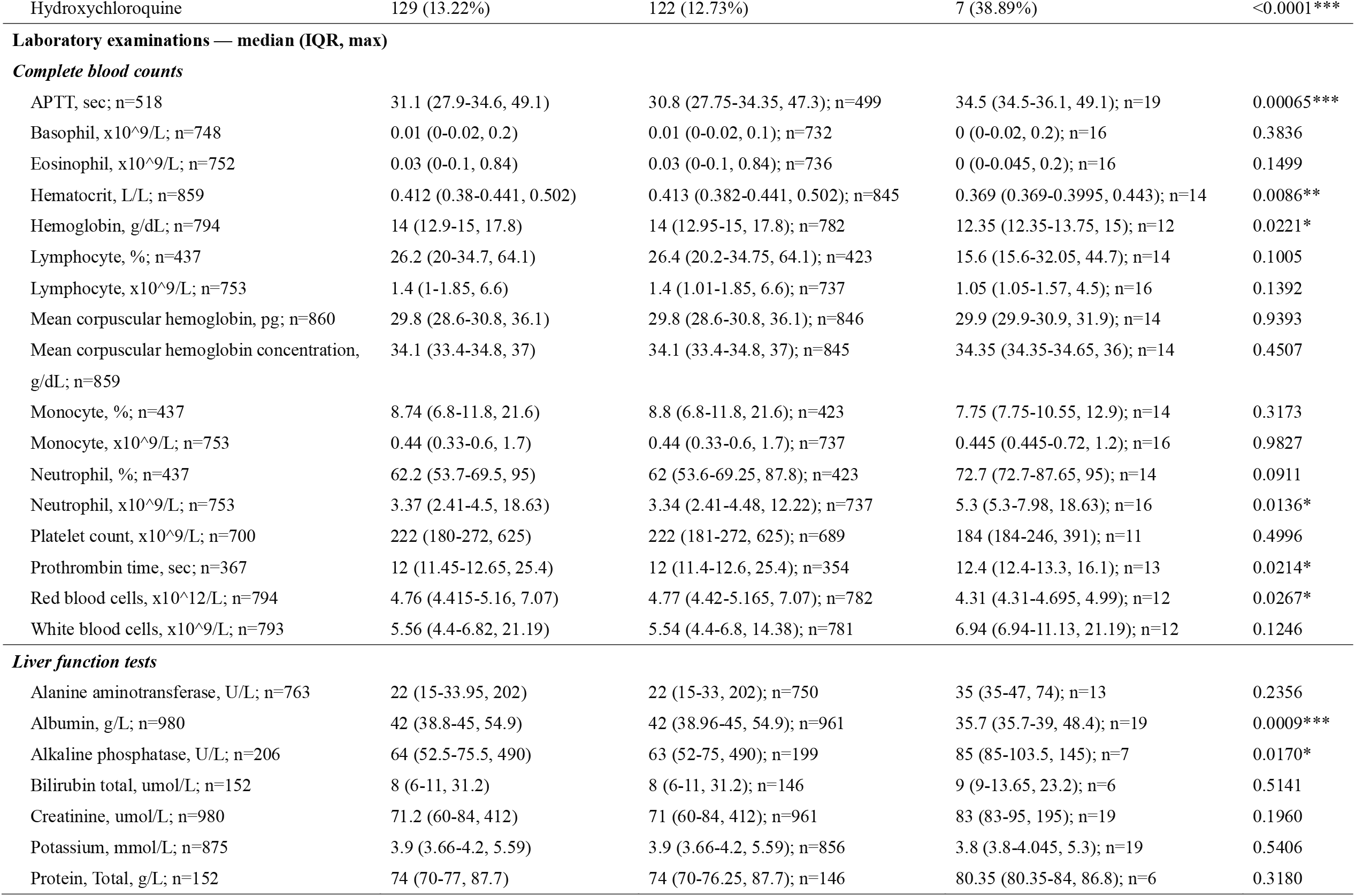
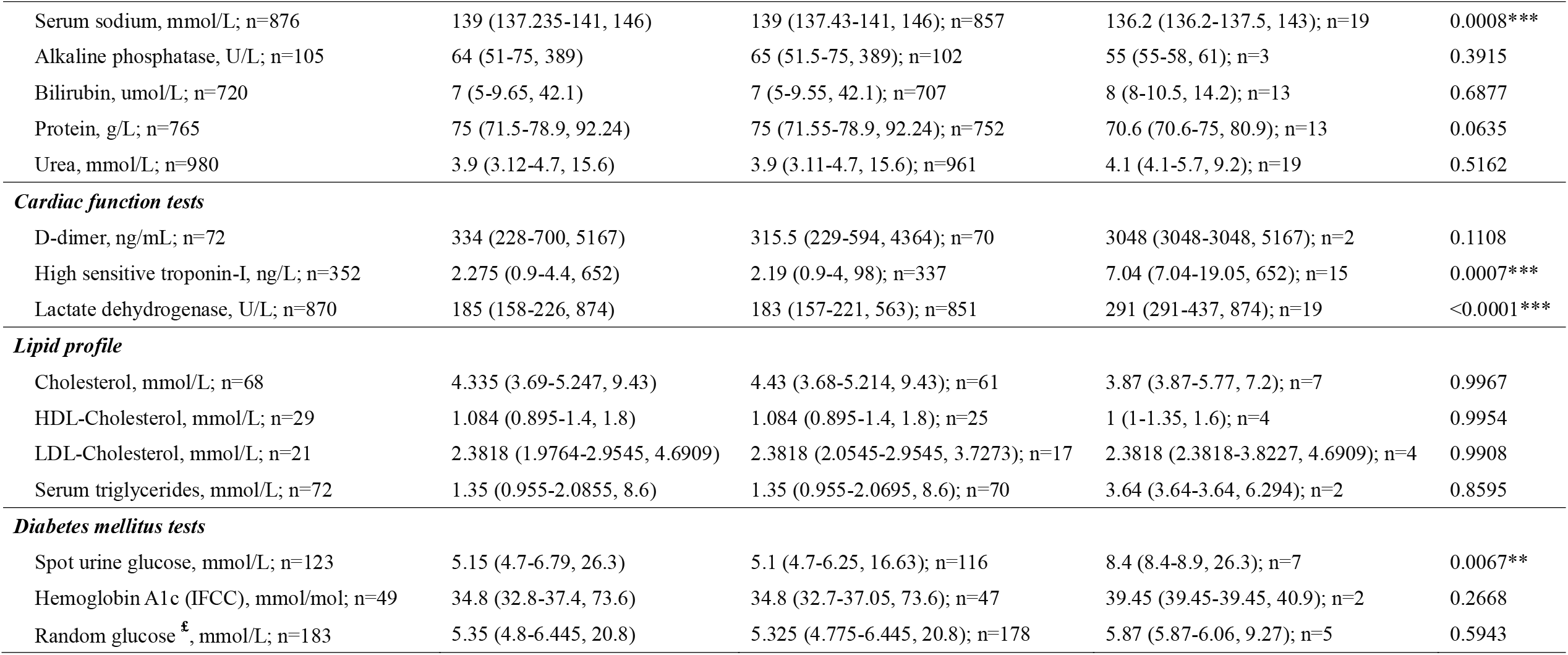
Demographic, epidemiological, clinical, medication, laboratory and ICU use outcome information collected from COVID-19 patients ICU=intensive unit care; COVID-19 = coronavirus disease 2019; APTT = Activated partial thromboplastin time; IQR = Interquartile range; * for p≤ 0.05, ** for p ≤ 0.01, *** for p ≤ 0.001 #: Descriptive statistics of individual comorbidities are included in supplementary material 1. *: Descriptive statistics of prescribed commodities and therapeutic classifications are included in supplementary material 2. Drug commodities are included in the Appendix. £: Random glucose test requires that blood sample is drawn at a laboratory at any time. Fasted or recently eaten will not affect the test.

| **Demographics** | All patients (n=1043) | Patients without ICU use (n=1034) | Patient with ICU use (n=19) | P value |
| --- | --- | --- | --- | --- |
| Age, year, n (IQR, max) | 34 (32-36, 96) | 34 (32-36, 96) | 59 (43-65, 93) | <0.0001*** |
| Sex, n (%) |  |  |  | 0.0040** |
| Male | 563 (53.98%) | 549 (53.61%) | 14 (73.68%) | - |
| Female | 480 (46.02 %) | 475 (46.39%) | 5 (26.32%) | - |
| **Epidemiological feature — n (%)** | All patients (n=1043) | Patients without ICU use (n=1034) | Patient with ICU use (n=19) | P value |
| Hong Kong island | 373 (35.67%) | 370 (36.13%) | 3 (15.79%) | 0.0014** |
| Kowloon | 212 (20.33%) | 209 (20.41%) | 3 (15.79%) | 0.6331 |
| New Territories | 398 (38.16%) | 386 (37.70%) | 12 (63.16%) | 0.0002*** |
| Others | 60 (5.75%) | 59 (5.76%) | 1 (5.26%) | 0.9847 |
| **Comorbidities<sup>#</sup> — n (%)** | All patients (n=535) | Patients without ICU use (n=524) | Patient with ICU use (n=11) | P value |
| Cardiovascular diseases | 10 (1.87%) | 3 (0.57%) | 7 (63.64%) | <0.0001*** |
| Lung and respiratory disease | 230 (42.99%) | 220 (41.98%) | 10 (90.91%) | <0.0001*** |
| Diabetes | 54 (10.09%) | 47 (8.97%) | 7 (63.64%) | <0.0001*** |
| Hypertension | 108 (20.19%) | 99 (18.89%) | 9 (81.82%) | <0.0001*** |
| Gastrointestinal diseases | 174 (32.52%) | 169 (32.25%) | 5 (45.45%) | 0.1050 |
| Chronic kidney diseases | 21 (3.93%) | 18 (3.44%) | 3 (27.27%) | <0.0001*** |
| **Drugs<sup>\*</sup> — n (%)** | All patients (n=976) | Patients without ICU use (n=958) | Patient with ICU use (n=18) | P value |
| ACEI or ARB | 184 (18.85%) | 170 (17.75%) | 14 (77.78%) | <0.0001*** |
| Steroids | 143 (14.65%) | 138 (14.41%) | 5 (27.78%) | 0.8831 |
| Kaletra | 593 (60.76%) | 577 (60.23%) | 16 (88.89%) | <0.0001*** |
| Ribavirin | 519 (53.18%) | 505 (52.71%) | 14 (77.78%) | 0.0002*** |
| Remdesivir | 24 (2.46%) | 23 (2.40%) | 1 (5.56%) | 0.3783 |
| Interferon beta | 317 (32.48%) | 313 (32.67%) | 4 (22.22%) | 0.1770 |
| Hydroxychloroquine | 129 (13.22%) | 122 (12.73%) | 7 (38.89%) | <0.0001*** |
| **Laboratory examinations — median (IQR, max)** |  |  |  |  |
| *Complete blood counts* |  |  |  |  |
| APTT, sec; n=518 | 31.1 (27.9-34.6, 49.1) | 30.8 (27.75-34.35, 47.3); n=499 | 34.5 (34.5-36.1, 49.1); n=19 | 0.00065*** |
| Basophil, x10<sup>9</sup>/L; n=748 | 0.01 (0-0.02, 0.2) | 0.01 (0-0.02, 0.1); n=732 | 0 (0-0.02, 0.2); n=16 | 0.3836 |
| Eosinophil, x10<sup>9</sup>/L; n=752 | 0.03 (0-0.1, 0.84) | 0.03 (0-0.1, 0.84); n=736 | 0 (0-0.045, 0.2); n=16 | 0.1499 |
| Hematocrit, L/L; n=859 | 0.412 (0.38-0.441, 0.502) | 0.413 (0.382-0.441, 0.502); n=845 | 0.369 (0.369-0.3995, 0.443); n=14 | 0.0086** |
| Hemoglobin, g/dL; n=794 | 14 (12.9-15, 17.8) | 14 (12.95-15, 17.8); n=782 | 12.35 (12.35-13.75, 15); n=12 | 0.0221* |
| Lymphocyte, %; n=437 | 26.2 (20-34.7, 64.1) | 26.4 (20.2-34.75, 64.1); n=423 | 15.6 (15.6-32.05, 44.7); n=14 | 0.1005 |
| Lymphocyte, x10<sup>9</sup>/L; n=753 | 1.4 (1-1.85, 6.6) | 1.4 (1.01-1.85, 6.6); n=737 | 1.05 (1.05-1.57, 4.5); n=16 | 0.1392 |
| Mean corpuscular hemoglobin, pg; n=860 | 29.8 (28.6-30.8, 36.1) | 29.8 (28.6-30.8, 36.1); n=846 | 29.9 (29.9-30.9, 31.9); n=14 | 0.9393 |
| Mean corpuscular hemoglobin concentration, g/dL; n=859 | 34.1 (33.4-34.8, 37) | 34.1 (33.4-34.8, 37); n=845 | 34.35 (34.35-34.65, 36); n=14 | 0.4507 |
| Monocyte, %; n=437 | 8.74 (6.8-11.8, 21.6) | 8.8 (6.8-11.8, 21.6); n=423 | 7.75 (7.75-10.55, 12.9); n=14 | 0.3173 |
| Monocyte, x10<sup>9</sup>/L; n=753 | 0.44 (0.33-0.6, 1.7) | 0.44 (0.33-0.6, 1.7); n=737 | 0.445 (0.445-0.72, 1.2); n=16 | 0.9827 |
| Neutrophil, %; n=437 | 62.2 (53.7-69.5, 95) | 62 (53.6-69.25, 87.8); n=423 | 72.7 (72.7-87.65, 95); n=14 | 0.0911 |
| Neutrophil, x10<sup>9</sup>/L; n=753 | 3.37 (2.41-4.5, 18.63) | 3.34 (2.41-4.48, 12.22); n=737 | 5.3 (5.3-7.98, 18.63); n=16 | 0.0136* |
| Platelet count, x10<sup>9</sup>/L; n=700 | 222 (180-272, 625) | 222 (181-272, 625); n=689 | 184 (184-246, 391); n=11 | 0.4996 |
| Prothrombin time, sec; n=367 | 12 (11.45-12.65, 25.4) | 12 (11.4-12.6, 25.4); n=354 | 12.4 (12.4-13.3, 16.1); n=13 | 0.0214* |
| Red blood cells, x10<sup>12</sup>/L; n=794 | 4.76 (4.415-5.16, 7.07) | 4.77 (4.42-5.165, 7.07); n=782 | 4.31 (4.31-4.695, 4.99); n=12 | 0.0267* |
| White blood cells, x10<sup>9</sup>/L; n=793 | 5.56 (4.4-6.82, 21.19) | 5.54 (4.4-6.8, 14.38); n=781 | 6.94 (6.94-11.13, 21.19); n=12 | 0.1246 |
| *Liver function tests* |  |  |  |  |
| Alanine aminotransferase, U/L; n=763 | 22 (15-33.95, 202) | 22 (15-33, 202); n=750 | 35 (35-47, 74); n=13 | 0.2356 |
| Albumin, g/L; n=980 | 42 (38.8-45, 54.9) | 42 (38.96-45, 54.9); n=961 | 35.7 (35.7-39, 48.4); n=19 | 0.0009*** |
| Alkaline phosphatase, U/L; n=206 | 64 (52.5-75.5, 490) | 63 (52-75, 490); n=199 | 85 (85-103.5, 145); n=7 | 0.0170* |
| Bilirubin total, umol/L; n=152 | 8 (6-11, 31.2) | 8 (6-11, 31.2); n=146 | 9 (9-13.65, 23.2); n=6 | 0.5141 |
| Creatinine, umol/L; n=980 | 71.2 (60-84, 412) | 71 (60-84, 412); n=961 | 83 (83-95, 195); n=19 | 0.1960 |
| Potassium, mmol/L; n=875 | 3.9 (3.66-4.2, 5.59) | 3.9 (3.66-4.2, 5.59); n=856 | 3.8 (3.8-4.045, 5.3); n=19 | 0.5406 |
| Protein, Total, g/L; n=152 | 74 (70-77, 87.7) | 74 (70-76.25, 87.7); n=146 | 80.35 (80.35-84, 86.8); n=6 | 0.3180 |
| Serum sodium, mmol/L; n=876 | 139 (137.235-141, 146) | 139 (137.43-141, 146); n=857 | 136.2 (136.2-137.5, 143); n=19 | 0.0008*** |
| Alkaline phosphatase, U/L; n=105 | 64 (51-75, 389) | 65 (51.5-75, 389); n=102 | 55 (55-58, 61); n=3 | 0.3915 |
| Bilirubin, umol/L; n=720 | 7 (5-9.65, 42.1) | 7 (5-9.55, 42.1); n=707 | 8 (8-10.5, 14.2); n=13 | 0.6877 |
| Protein, g/L; n=765 | 75 (71.5-78.9, 92.24) | 75 (71.55-78.9, 92.24); n=752 | 70.6 (70.6-75, 80.9); n=13 | 0.0635 |
| Urea, mmol/L; n=980 | 3.9 (3.12-4.7, 15.6) | 3.9 (3.11-4.7, 15.6); n=961 | 4.1 (4.1-5.7, 9.2); n=19 | 0.5162 |
| ***Cardiac function tests*** |  |  |  |  |
| D-dimer, ng/mL; n=72 | 334 (228-700, 5167) | 315.5 (229-594, 4364); n=70 | 3048 (3048-3048, 5167); n=2 | 0.1108 |
| High sensitive troponin-I, ng/L; n=352 | 2.275 (0.9-4.4, 652) | 2.19 (0.9-4, 98); n=337 | 7.04 (7.04-19.05, 652); n=15 | 0.0007*** |
| Lactate dehydrogenase, U/L; n=870 | 185 (158-226, 874) | 183 (157-221, 563); n=851 | 291 (291-437, 874); n=19 | <0.0001*** |
| ***Lipid profile*** |  |  |  |  |
| Cholesterol, mmol/L; n=68 | 4.335 (3.69-5.247, 9.43) | 4.43 (3.68-5.214, 9.43); n=61 | 3.87 (3.87-5.77, 7.2); n=7 | 0.9967 |
| HDL-Cholesterol, mmol/L; n=29 | 1.084 (0.895-1.4, 1.8) | 1.084 (0.895-1.4, 1.8); n=25 | 1 (1-1.35, 1.6); n=4 | 0.9954 |
| LDL-Cholesterol, mmol/L; n=21 | 2.3818 (1.9764-2.9545, 4.6909) | 2.3818 (2.0545-2.9545, 3.7273); n=17 | 2.3818 (2.3818-3.8227, 4.6909); n=4 | 0.9908 |
| Serum triglycerides, mmol/L; n=72 | 1.35 (0.955-2.0855, 8.6) | 1.35 (0.955-2.0695, 8.6); n=70 | 3.64 (3.64-3.64, 6.294); n=2 | 0.8595 |
| ***Diabetes mellitus tests*** |  |  |  |  |
| Spot urine glucose, mmol/L; n=123 | 5.15 (4.7-6.79, 26.3) | 5.1 (4.7-6.25, 16.63); n=116 | 8.4 (8.4-8.9, 26.3); n=7 | 0.0067** |
| Hemoglobin A1c (IFCC), mmol/mol; n=49 | 34.8 (32.8-37.4, 73.6) | 34.8 (32.7-37.05, 73.6); n=47 | 39.45 (39.45-39.45, 40.9); n=2 | 0.2668 |
| Random glucose <sup>‡</sup>, mmol/L; n=183 | 5.35 (4.8-6.445, 20.8) | 5.325 (4.775-6.445, 20.8); n=178 | 5.87 (5.87-6.06, 9.27); n=5 | 0.5943 |

**Table 3.**
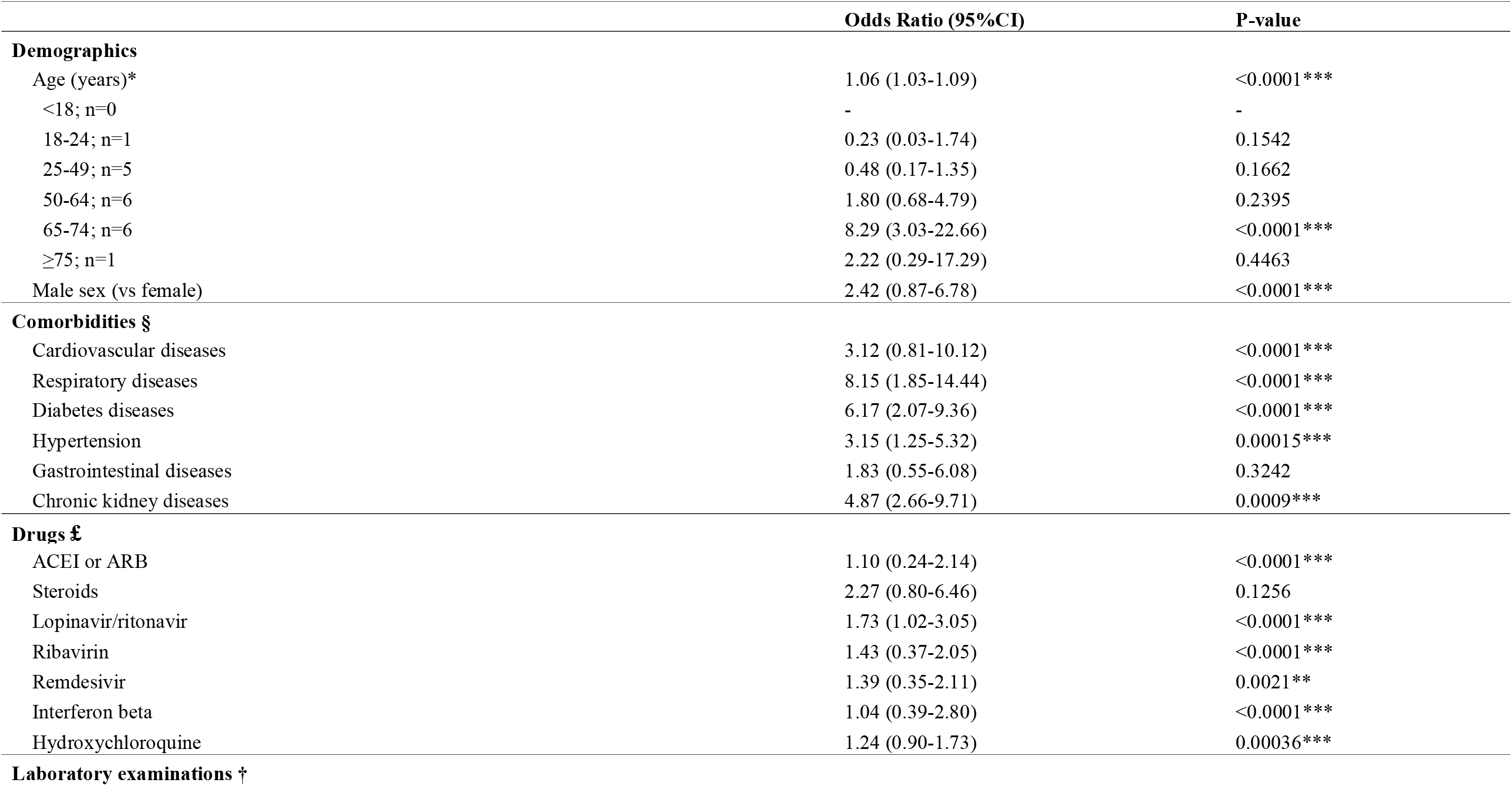
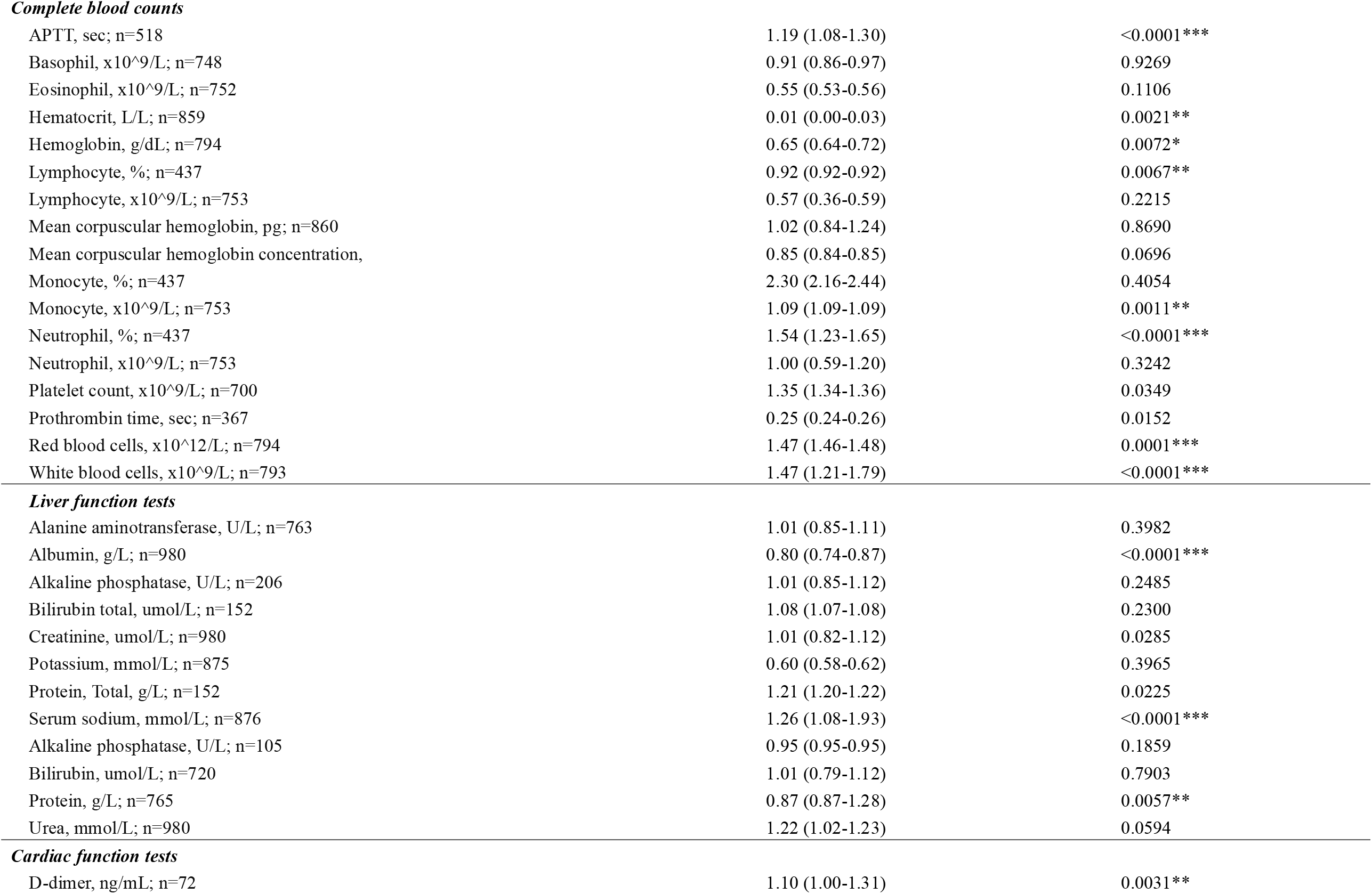
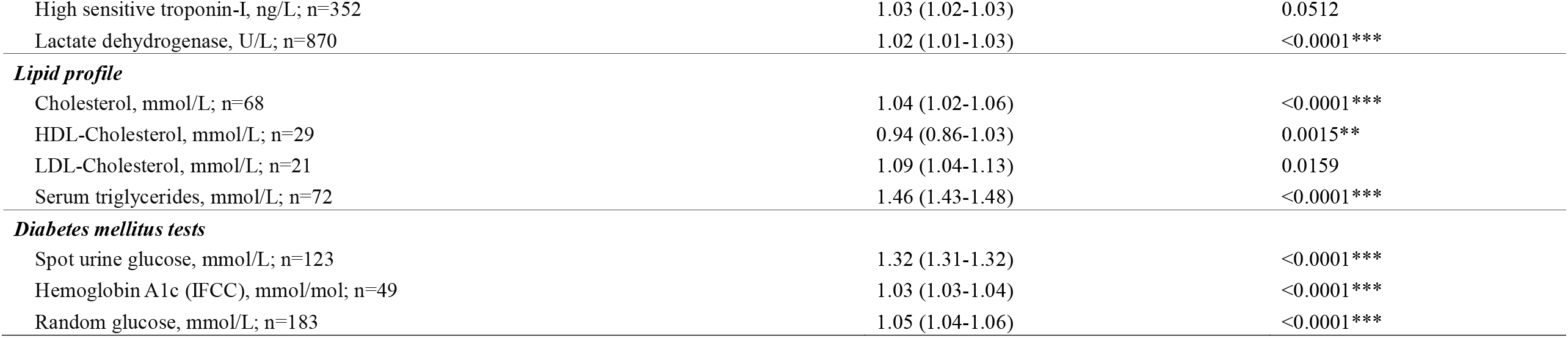
Predictors of ICU admission in COVID-19 patients. ICU=intensive unit care; COVID-19 = coronavirus disease 2019; APTT=Activated partial thromboplastin time; IQR=Interquartile range; * for p≤ 0.05, ** for p ≤ 0.01, *** for p ≤ 0.001 All analyses were adjusted for age, sex, comorbidities and residence districts unless otherwise specified. *: Adjusted for sex, comorbidities, and residence districts. §: Adjusted for sex, and residence districts. £: Adjusted for sex, comorbidities. Univariate analysis results of individual drugs are included in in supplementary material 3. †: Adjusted for sex, comorbidities, and residence districts.

|  | Odds Ratio (95%CI) | P-value |
| --- | --- | --- |
| **Demographics** |  |  |
| Age (years)* | 1.06 (1.03-1.09) | <0.0001*** |
| <18; n=0 | - | - |
| 18-24; n=1 | 0.23 (0.03-1.74) | 0.1542 |
| 25-49; n=5 | 0.48 (0.17-1.35) | 0.1662 |
| 50-64; n=6 | 1.80 (0.68-4.79) | 0.2395 |
| 65-74; n=6 | 8.29 (3.03-22.66) | <0.0001*** |
| ≥75; n=1 | 2.22 (0.29-17.29) | 0.4463 |
| Male sex (vs female) | 2.42 (0.87-6.78) | <0.0001*** |
| **Comorbidities §** |  |  |
| Cardiovascular diseases | 3.12 (0.81-10.12) | <0.0001*** |
| Respiratory diseases | 8.15 (1.85-14.44) | <0.0001*** |
| Diabetes diseases | 6.17 (2.07-9.36) | <0.0001*** |
| Hypertension | 3.15 (1.25-5.32) | 0.00015*** |
| Gastrointestinal diseases | 1.83 (0.55-6.08) | 0.3242 |
| Chronic kidney diseases | 4.87 (2.66-9.71) | 0.0009*** |
| **Drugs £** |  |  |
| ACEI or ARB | 1.10 (0.24-2.14) | <0.0001*** |
| Steroids | 2.27 (0.80-6.46) | 0.1256 |
| Lopinavir/ritonavir | 1.73 (1.02-3.05) | <0.0001*** |
| Ribavirin | 1.43 (0.37-2.05) | <0.0001*** |
| Remdesivir | 1.39 (0.35-2.11) | 0.0021** |
| Interferon beta | 1.04 (0.39-2.80) | <0.0001*** |
| Hydroxychloroquine | 1.24 (0.90-1.73) | 0.00036*** |
| **Laboratory examinations †** |  |  |
| ***Complete blood counts*** |  |  |
| APTT, sec; n=518 | 1.19 (1.08-1.30) | <0.0001*** |
| Basophil, x10<sup>9</sup>/L; n=748 | 0.91 (0.86-0.97) | 0.9269 |
| Eosinophil, x10<sup>9</sup>/L; n=752 | 0.55 (0.53-0.56) | 0.1106 |
| Hematocrit, L/L; n=859 | 0.01 (0.00-0.03) | 0.0021** |
| Hemoglobin, g/dL; n=794 | 0.65 (0.64-0.72) | 0.0072* |
| Lymphocyte, %; n=437 | 0.92 (0.92-0.92) | 0.0067** |
| Lymphocyte, x10<sup>9</sup>/L; n=753 | 0.57 (0.36-0.59) | 0.2215 |
| Mean corpuscular hemoglobin, pg; n=860 | 1.02 (0.84-1.24) | 0.8690 |
| Mean corpuscular hemoglobin concentration, | 0.85 (0.84-0.85) | 0.0696 |
| Monocyte, %; n=437 | 2.30 (2.16-2.44) | 0.4054 |
| Monocyte, x10<sup>9</sup>/L; n=753 | 1.09 (1.09-1.09) | 0.0011** |
| Neutrophil, %; n=437 | 1.54 (1.23-1.65) | <0.0001*** |
| Neutrophil, x10<sup>9</sup>/L; n=753 | 1.00 (0.59-1.20) | 0.3242 |
| Platelet count, x10<sup>9</sup>/L; n=700 | 1.35 (1.34-1.36) | 0.0349 |
| Prothrombin time, sec; n=367 | 0.25 (0.24-0.26) | 0.0152 |
| Red blood cells, x10<sup>12</sup>/L; n=794 | 1.47 (1.46-1.48) | 0.0001*** |
| White blood cells, x10<sup>9</sup>/L; n=793 | 1.47 (1.21-1.79) | <0.0001*** |
| ***Liver function tests*** |  |  |
| Alanine aminotransferase, U/L; n=763 | 1.01 (0.85-1.11) | 0.3982 |
| Albumin, g/L; n=980 | 0.80 (0.74-0.87) | <0.0001*** |
| Alkaline phosphatase, U/L; n=206 | 1.01 (0.85-1.12) | 0.2485 |
| Bilirubin total, umol/L; n=152 | 1.08 (1.07-1.08) | 0.2300 |
| Creatinine, umol/L; n=980 | 1.01 (0.82-1.12) | 0.0285 |
| Potassium, mmol/L; n=875 | 0.60 (0.58-0.62) | 0.3965 |
| Protein, Total, g/L; n=152 | 1.21 (1.20-1.22) | 0.0225 |
| Serum sodium, mmol/L; n=876 | 1.26 (1.08-1.93) | <0.0001*** |
| Alkaline phosphatase, U/L; n=105 | 0.95 (0.95-0.95) | 0.1859 |
| Bilirubin, umol/L; n=720 | 1.01 (0.79-1.12) | 0.7903 |
| Protein, g/L; n=765 | 0.87 (0.87-1.28) | 0.0057** |
| Urea, mmol/L; n=980 | 1.22 (1.02-1.23) | 0.0594 |
| ***Cardiac function tests*** |  |  |
| D-dimer, ng/mL; n=72 | 1.10 (1.00-1.31) | 0.0031** |
| High sensitive troponin-I, ng/L; n=352 | 1.03 (1.02-1.03) | 0.0512 |
| Lactate dehydrogenase, U/L; n=870 | 1.02 (1.01-1.03) | <0.0001*** |
| ***Lipid profile*** |  |  |
| Cholesterol, mmol/L; n=68 | 1.04 (1.02-1.06) | <0.0001*** |
| HDL-Cholesterol, mmol/L; n=29 | 0.94 (0.86-1.03) | 0.0015** |
| LDL-Cholesterol, mmol/L; n=21 | 1.09 (1.04-1.13) | 0.0159 |
| Serum triglycerides, mmol/L; n=72 | 1.46 (1.43-1.48) | <0.0001*** |
| ***Diabetes mellitus tests*** |  |  |
| Spot urine glucose, mmol/L; n=123 | 1.32 (1.31-1.32) | <0.0001*** |
| Hemoglobin A1c (IFCC), mmol/mol; n=49 | 1.03 (1.03-1.04) | <0.0001*** |
| Random glucose, mmol/L; n=183 | 1.05 (1.04-1.06) | <0.0001*** |

1. Demographic features: Age (OR: 1.06 [1.03 -1.09], p<0.0001) and male (OR: 2.42 [0.87-6.78], p<0.0001).
2. Comorbidities: cardiovascular diseases (OR: 3.12 [0.81-10.12], p<0.0001), respiratory diseases (OR: 8.15 [1.85-14.44], p<0.0001), diabetes (OR: 6.17 [2.07-9.36], p<0.0001), hypertension (OR: 3.15 [1.25-5.32], p<0.0001) and chronic kidney diseases (OR: 4.87 [2.66-9.71], p=0.0009). The analyses demonstrate the importance of baseline comorbidities in affecting the prognosis of patients with COVID-19.
3. Drugs: ACEI or ARB (OR: 1.10 [0.24-2.14], p<0.0001), lopinavir/ritonavir (OR: 1.73 [1.02-3.05], p<0.0001), ribavirin (OR: 1.43 [0.37-2.05], p<0.0001), remdesivir (OR: 1.04 [0.39-2.80], p<0.0001), interferon beta (OR: 1.04 [0.39-2.80], p<0.0001) and hydroxychloroquine (OR: 1.24 [0.90-1.73], p=0.00036).
4. Biochemical markers: APTT (OR: 1.19 [1.08-1.30], p=0.0003), neutrophil count (OR: 1.54 [1.53-1.55], p<0.0001), red blood cells (OR: 1.47 [1.46-1.48], p<0.0001), white blood cells (OR: 1.47 [1.21-1.79], p<0.0001), albumin (OR: 0.80 [0.74-0.87], p<0.0001), serum sodium (OR: 1.26 [1.08-1.93], p<0.0001), lactate dehydrogenase (OR: 1.01 [0.85-1.12], p<0.0001), total cholesterol (OR: 1.04 [1.02-1.06], p<0.0001), spot urine glucose (OR: 1.32 [1.31-1.32], p<0.0001), hemoglobin A1c (OR: 1.03 [1.03-1.04]<0.0001), random glucose (OR: 1.05 [1.04-1.06], p<0.0001), serum triglycerides (OR: 1.46 [1.43-1.48], p<0.0001).

### Main and Hidden Interaction Effects

The EBM model was employed to distinguish patients in need for ICU admission by accurately uncovering the main and hidden interaction effects. This utilized different data modalities such as demographics, comorbidities and multiple laboratory results. Significant variables identified by univariate logistic regression were entered into the EBM model, which will deal with the trade-off between having a minimal number of predictors and the capacity of good model prediction, therefore avoiding overfitting. The cohort is randomly classified into training and validation datasets with an 80:20 split. The obtained importance rankings of significant predictors for ICU admission are shown in **Figure 5**. Red blood cells, APTT, sex, age and white blood cells are the five most informative parameters in predicting ICU admission, followed by hypertension, serum sodium, serum albumin, serum triglycerides, and respiratory disease. Significant predictors for ICU admission identification are provided in **Figure 6**. We can observe that the following combination of patient characteristics predicts a higher likelihood for ICU admission: 1) male patients with lower level of red blood cells, 2) older patients with lower level of red blood cells, 3) patients with both lower levels of red blood cells and albumin or sodium, 4) patients with longer APTT and lower level of red blood cells. Important hidden pair-wise interaction effects are shown in **Figure 7**, where green or yellow zones with larger values indicate higher probability of ICU admission that can be predicted by examining the pair-wise variable interactions. We can observe from the plots of interaction effects that 1) male with lower red blood cells, (2) older age with lower red blood cells, 3) lower albumin level and lower red blood cells, 4) lower sodium level and lower red blood cells, 5) older age and prolonged APTT, 6) lower red bold cells level and higher white blood cells level, 7) lower red blood cells level and prolonged APTT, 8) older age and higher level white predicts higher probability of ICU admission.

**Figure 5.**
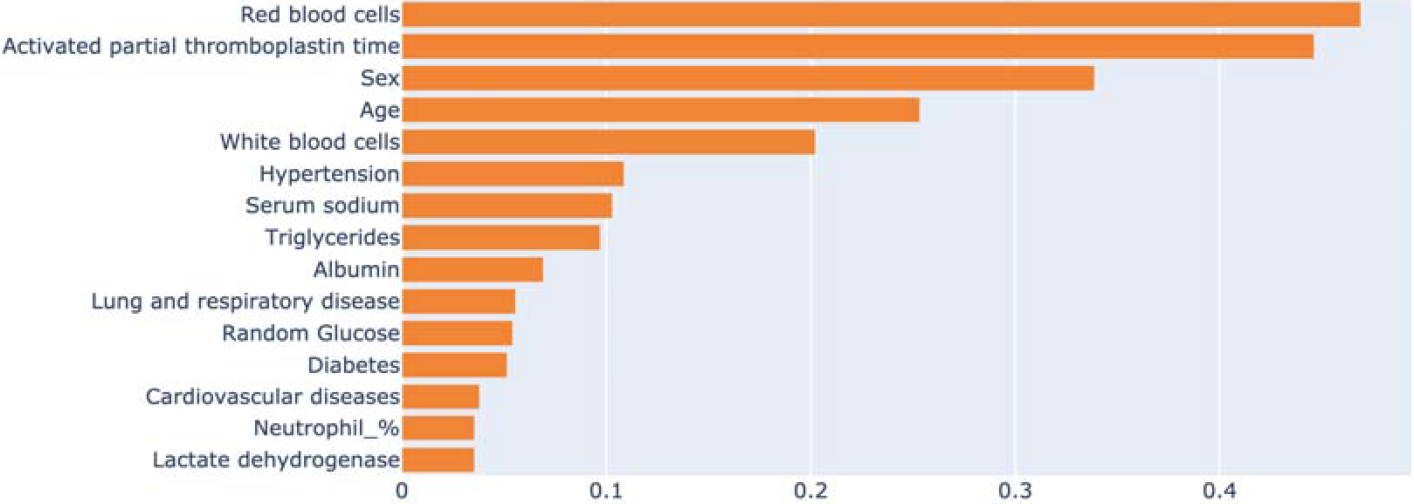
Importance ranking of significant univariable characteristics for ICU identification.

**Figure 6.**
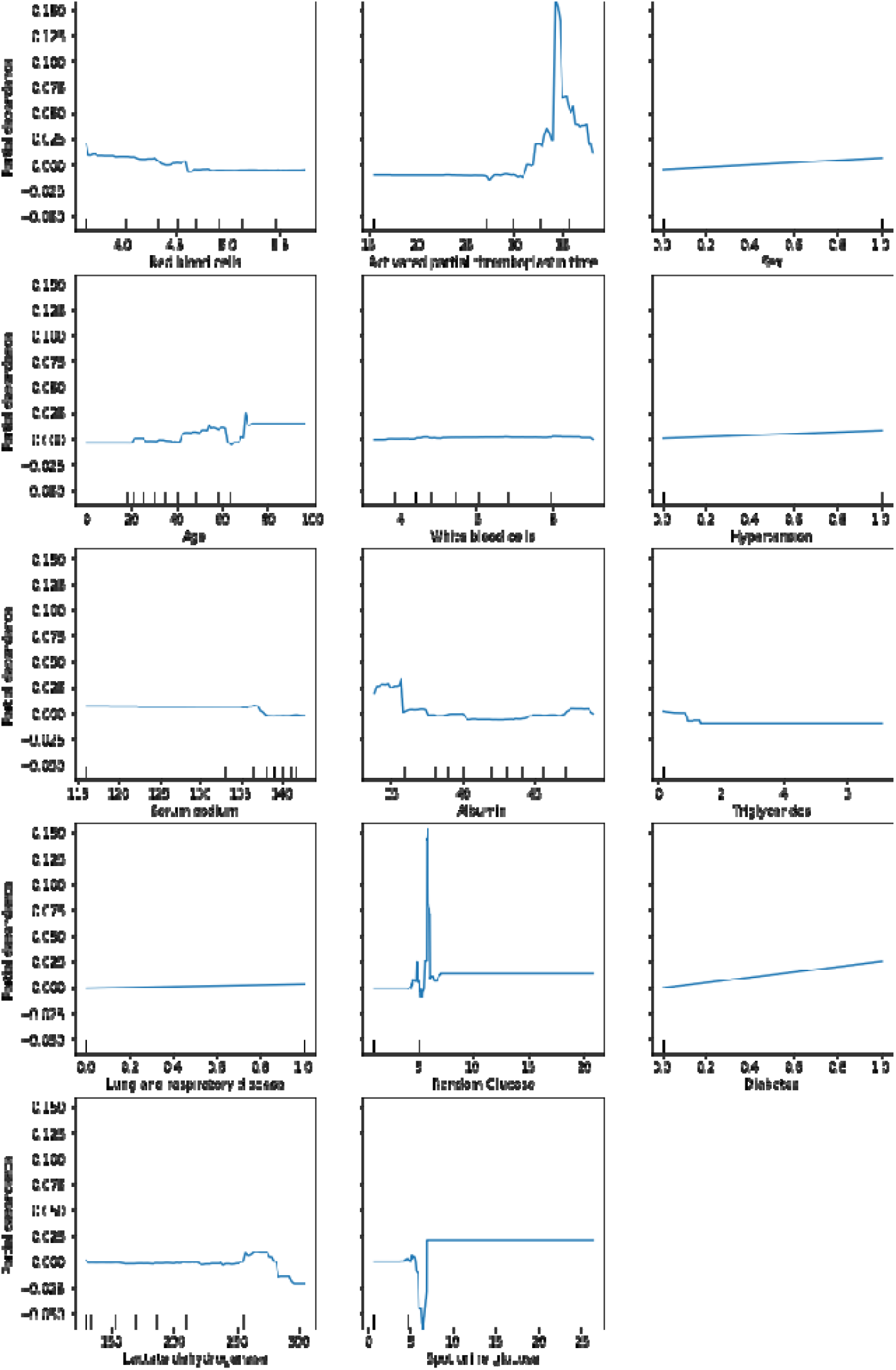
Changing effects of significant predictors on ICU use identification.

**Figure 7.**
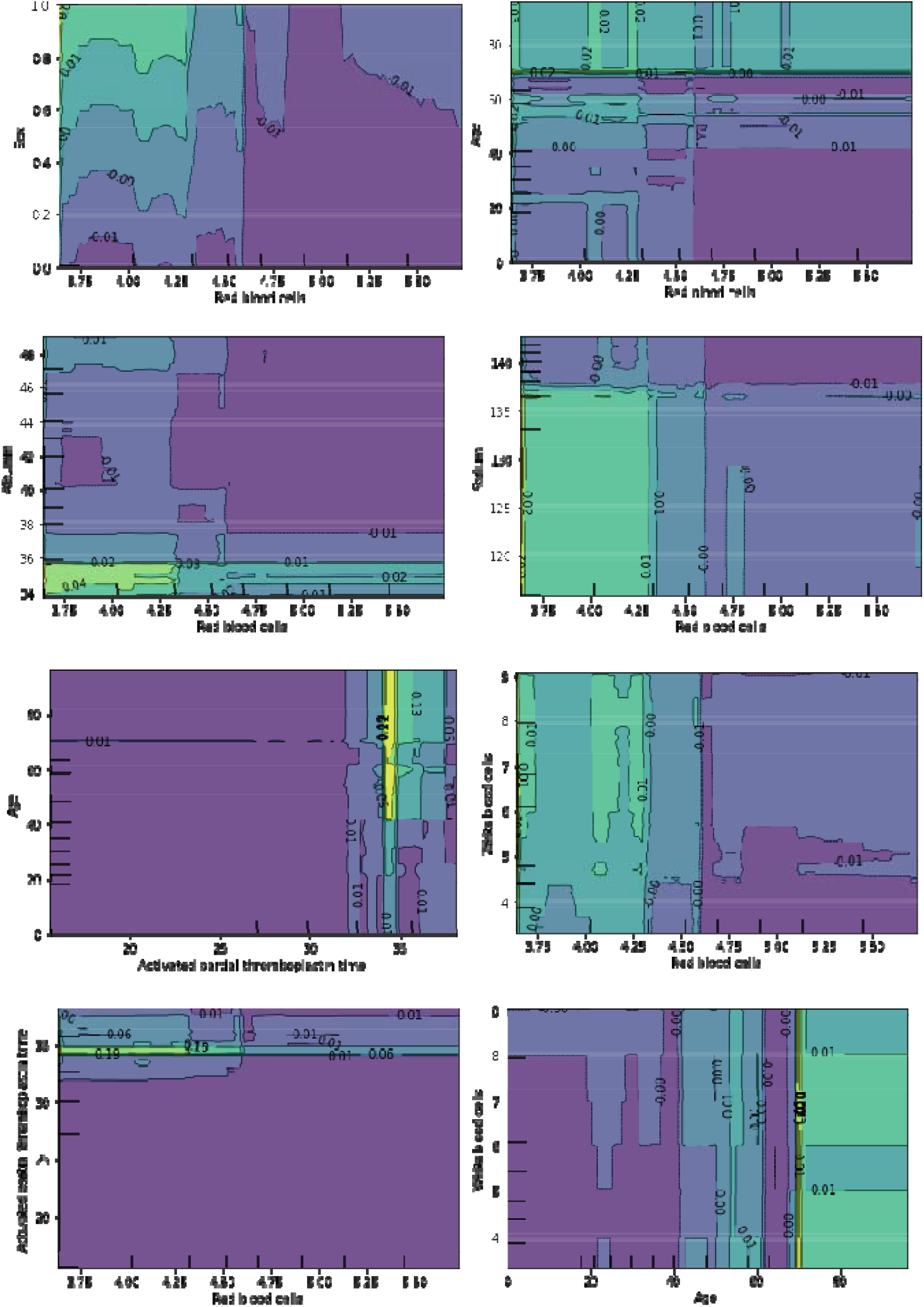
Interaction effects of important pairwise univariable characteristics for ICU identification.

EBM can provide predictions on individual cases. For example, a randomly selected patient (male, 69 years old) with ICU admission has the characteristics as shown in **Figure 8**. He has prior comorbidities of cardiovascular, chronic kidney, hypertension, diabetes and lung and respiratory diseases. EBM predicts that he needs ICU attention with 72% probability, based on his characteristics of prior cardiovascular disease, white blood cells at 12.43 (x10^9/L), lactate dehydrogenase level at 390 (U/L), APTT at 33.70 (sec), prior comorbidities of hypertension and diabetes, and others. But his characteristic of triglycerides at 6.29 provide non-supportive information to the prediction outcome. By contrast, a randomly selected patient (female, 54 years old) who did not require ICU admission is exemplified in **Figure 9**. EBM accurately predicted that she doesn’t need ICU admission. Local explanations provided by EBM can provide precise ICU admission predictions based on patient’s main characteristics in a user-friendly visualization way for practical clinical use.

**Figure 8.**
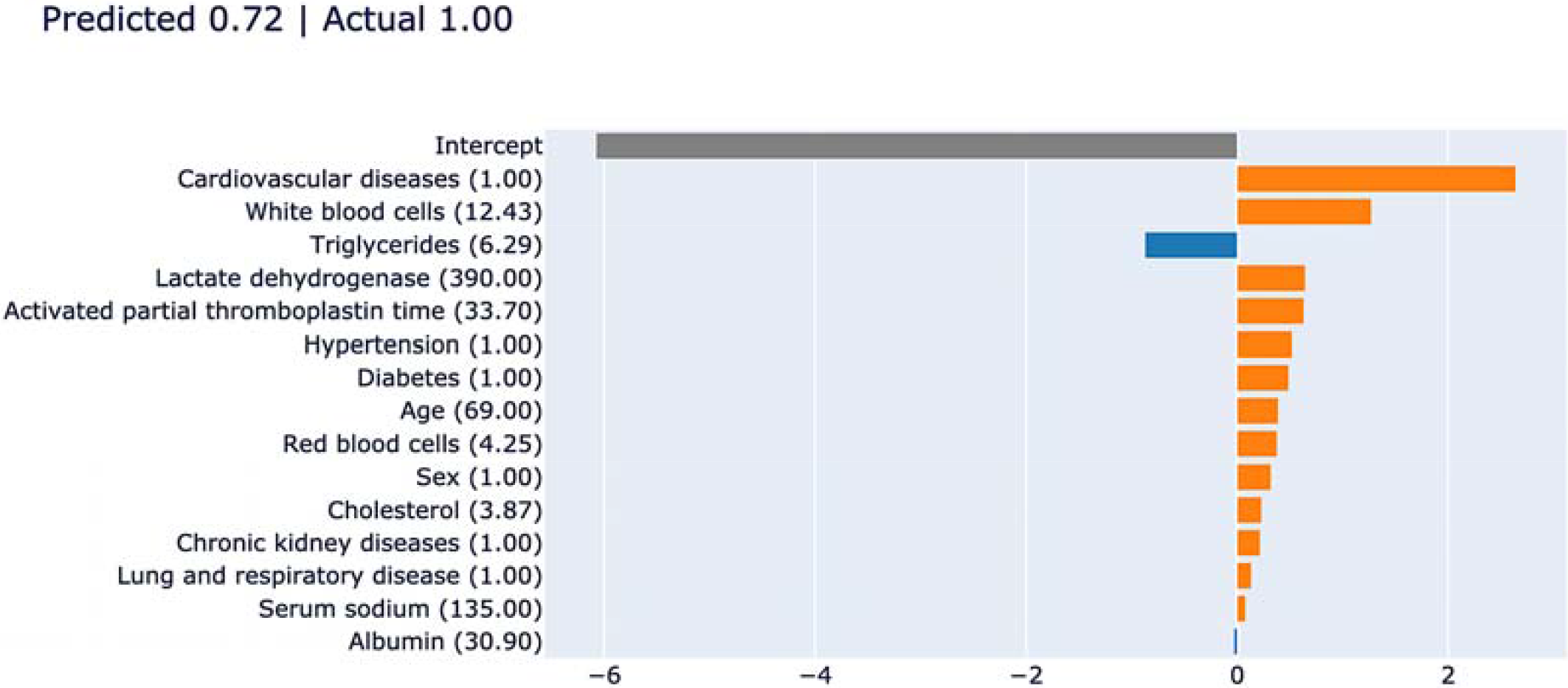
Local explanation for a random patient with ICU admission.

**Figure 9.**
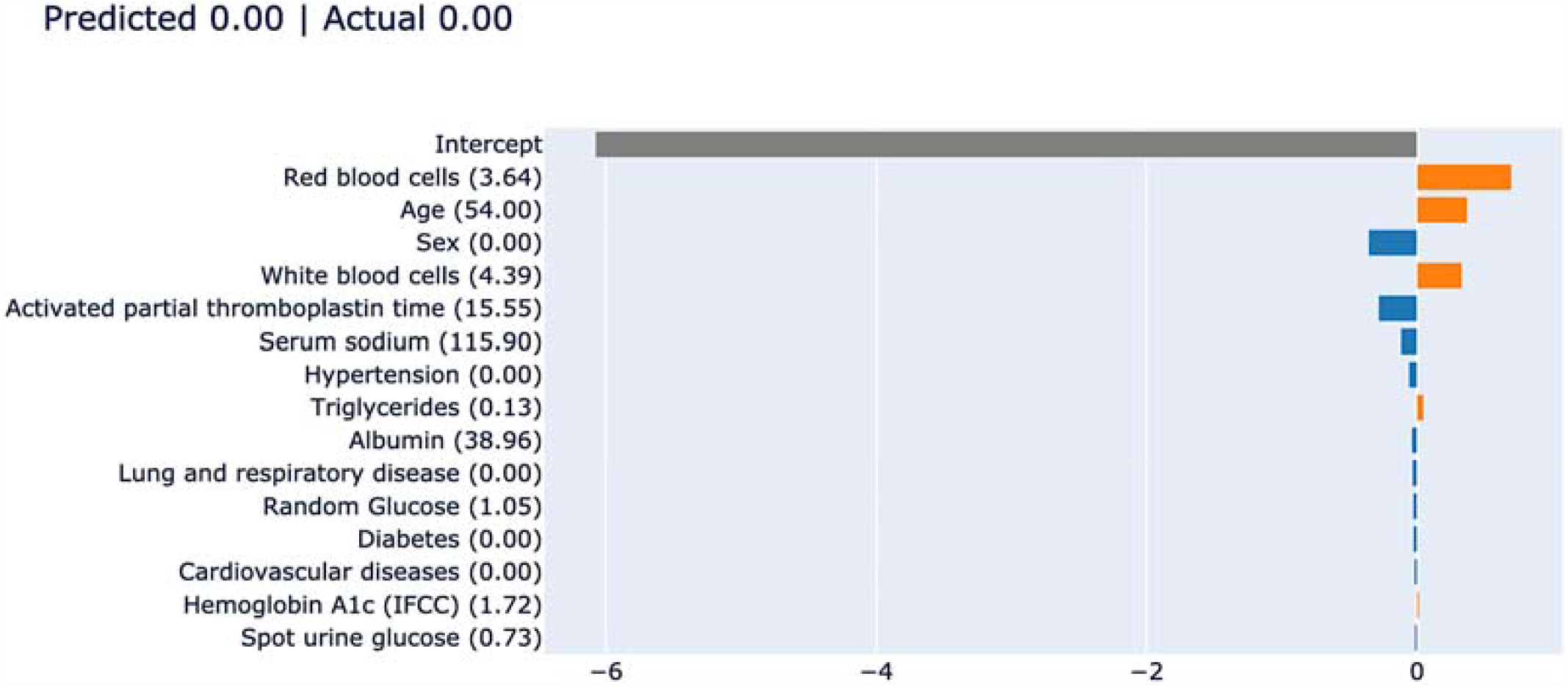
Local explanation for a random patient without ICU admission.

The five-fold cross validation performance of EBM was compared with baseline models including XGBoost, LightGBM, random forests, and multivariate logistic regression, as shown in **Table 4**. EBM outperforms all baseline models according to evaluation metrics of precision, recall, F1 score, and area under the curve (AUC) of the receiving operating characteristics (ROC) curve.

**Table 4.** Performance analysis of EBM over baseline models

|  | **Precision** | **Recall** | **F1 Score** | **AUC** |
| --- | --- | --- | --- | --- |
| EBM | 0.9117 | 0.9263 | 0.9189 | 0.9231 |
| XGBoost | 0.9074 | 0.8976 | 0.9025 | 0.8982 |
| LightGBM | 0.8337 | 0.8501 | 0.8418 | 0.8040 |
| Random forest | 0.8188 | 0.8215 | 0.8201 | 0.8250 |
| Logistic regression | 0.8337 | 0.8041 | 0.8186 | 0.8310 |

## Discussion

The main findings of this territory-wide retrospective cohort study are twofold: (1) Significant predictors of ICU admission were older age, male sex, prior coronary artery disease, respiratory diseases, diabetes, hypertension and chronic kidney disease, and activated partial thromboplastin time, red cell count, white cell count, albumin and sodium; (2) A tree-based interpretable machine learning model identified most informative characteristics and hidden interactions that can predict ICU admission. These interacting factors were low red cells with 1) male, 2) older age, 3) low albumin, 4) low sodium or 5) prolonged APTT around 33 seconds.

Prior studies have reported that patients with pre-existing medical comorbidities have a poorer prognosis in not only COVID-19 but also other infectious diseases such as SARS-CoV and MERS ^14, 15^. In COVID-19, hypertension, diabetes, coronary heart disease, chronic kidney disease, cerebrovascular disease, hepatitis, and chronic obstructive pulmonary disease (COPD) have been identified as predictors of disease severity and mortality in COVID-19 ^16, 17^. In this study, we confirm that these comorbidities are predictive of ICU utilization and provide a simple clinical approach to quantify the initial risk of ICU admission precisely and quickly. Furthermore, various laboratory markers have been shown to predict adverse outcomes. Our study found that prolonged APTT and raised D-dimer, reflecting coagulopathy, was predictive of ICU admission. Other significant predictors were neutrophil count (inflammation), red cell count (oxygen carrying capacity), albumin (nutritional status), sodium (electrolyte homeostasis) and lactate dehydrogenase (tissue damage). Troponin was borderline significant, reflecting that myocardial damage is an important determinant of ICU use.

We further illustrate the novel findings that interacting factors between low red cell count and basic demographics such as gender and age, or laboratory findings such as albumin, sodium and APTT are also important determinants. Older patients with laboratory examinations of lower red cells, lower albumin, lower sodium and prolonged APTT are subject to high ICU admission risk. Red cells, albumin, sodium and APTT can be easily collected in any hospital. In crowded hospitals with limited medical resources, this simple model can help to quickly prioritize patients for ICU attention.

The optimum medication regimen for COVID-19 is yet to be determined. However, small scale observational studies or trials have suggested the use of antivirals ^18^, antimalarials ^19^, interferons ^20^, anticoagulants ^21^ and antibodies ^22^, though not all have been shown to be beneficial in larger clinical trials ^23^. A better understanding of the pathophysiological mechanisms underlying COVID-19 will enable better treatment strategies to be devised ^24^. In our study, the anti-viral drug lopinavir/ritonavir (Kaletra) was the commonest prescribed drug, followed by ribavirin, interferon-beta, ACEIs/ARBs, steroids, hydroxychloroquine and the antiviral remdesivir. We found that these medications were more frequently prescribed in patients requiring ICU compared to those without. This may reflect the increased severity of cases in which clinicians were more likely to prescribe a cocktail of drugs.

## Conclusion

In summary, this study has identified important univariable and interaction effects informing intensive care admission in patients hospitalized with COVID-19. Significant univariable predictors of ICU admission include older age, male sex, prior coronary artery disease, respiratory diseases, diabetes, hypertension and chronic kidney disease, and activated partial thromboplastin time, red cell count, white cell count, albumin and serum sodium. A tree-based interpretable machine learning model identified most informative characteristics and hidden interactions (i.e., low red cells with male, older age, low albumin, low sodium or prolonged APTT) for COVID-19 prognostic ICU admission prediction. The tree-based machine learning model outperforms several baselines, enabling early detection of ICU admission, efficient healthcare resource utilization, and potentially mortality reduction of hospitalized patients with COVID-19.

## Data Availability

Available upon request

